# A SIRD model applied to COVID-19 dynamics and intervention strategies during the first wave in Kenya

**DOI:** 10.1101/2021.03.17.21253626

**Authors:** Wandera Ogana, Victor Ogesa Juma, Wallace D. Bulimo

## Abstract

The first case of COVID-19 was reported in Kenya in March 2020 and soon after non-pharmaceutical interventions (NPIs) were established to control the spread of the disease. The NPIs consisted, and continue to consist, of mitigation measures followed by a period of relaxation of some of the measures. In this paper, we use a deterministic mathematical model to analyze the dynamics of the disease, during the first wave, and relate it to the intervention measures. In the process, we develop a new method for estimating the disease parameters. Our solutions yield a basic reproduction number, R_0_ = 2.76, which is consistent with other solutions. The results further show that the initial mitigation reduced disease transmission by 40% while the subsequent relaxation increased transmission by 25%. We also propose a mathematical model on how interventions of known magnitudes collectively affect disease transmission rates. The modelled positivity rate curve compares well with observations. If interventions of unknown magnitudes have occurred, and data is available on the positivity rate, we use the method of planar envelopes around a curve to deduce the modelled positivity rate and the magnitudes of the interventions. Our solutions deduce mitigation and relaxation effects of 42.5% and 26%, respectively; these percentages are close to values obtained by the solution of the SIRD system. Our methods so far apply to a single wave; there is a need to investigate the possibility of extending them to handle multiple waves.

## 1 Introduction

Coronavirus Disease of 2019 (COVID-19) is the disease caused by the novel coronavirus that appeared in Wuhan, China, in December 2019. The disease has since spread to all parts of the world and resulted in 103,513,141 confirmed infections with 2,237,247 deaths by 31st January 2021 [1]. In Africa, the first reported case of COVID-19 was on 14th February 2020, in Egypt [2]. It has since afflicted 47all countries in the continent and led to 3,585,676 confirmed infections with 91,079 deaths to 31st January 2021 [1]. To control the spread of the disease, countries have introduced several non-pharmaceutical interventions (NPIs) in addition to strengthening health facilities and treatment regimes. The disease has wreaked havoc on the economies of most countries and transformed people’s lifestyles permanently.

We briefly present pertinent biological information about COVID-19 and a related disease, influenza, popularly known as flu. Our reference to influenza here will mean seasonal and not pandemic influenza. In the early days of COVID-19 many people confused the disease with flu since they both yield almost identical symptoms. On 11th February 2020, the virus which causes COVID-19 was named “severe acute respiratory syndrome coronavirus 2 (SARS-CoV-2” by the International Committee on Taxonomy of Viruses (ICTV) [3]. The flu is caused by influenza virus types A, B and C. SARS-CoV-2 and influenza viruses use different receptors to enter the host cell; the former uses the spike (S) protein for entry [4] while the latter uses the hemagglutinin (HA) protein [5]. Both viruses are spread through droplets released from the nose and mouth of an infected individual as they cough or sneeze [6]. Nevertheless, unlike influenza, it has been shown that COVID-19 may also be spread by the long-range airborne route at greater distances [7]. A critical determinant of the infectivity of these viruses is the concept of reproduction number, *R*_0_, which represents the degree of transmissibility of the virus and provides a representation of how many people can be infected by one person infected with the virus, in a population where everyone is susceptible to the disease. In the early stages of the epidemic, the *R*_0_ for COVID-19 was estimated to be 2.2 – 2.5 [8]; since then most of the published estimates for *R*_0_ lie in the range 2 – 3, with some as high as 6 and others as low as 1.9 [9,10]. For seasonal influenza, studies yielded *R*_0_ in the range 1.1 – 1.5 [11], suggesting that COVID-19 is more easily spread than seasonal influenza. For both flu and COVID-19, it is possible to spread the virus at least one day before experiencing any symptoms. Once an individual has flu, the person may be contagious for 5 – 7 days while for COVID-19 the person could be contagious for 10 – 14 days; the number of days of staying contagious will depend on the age of the patient, severity of the disease and whether there are other underlying medical conditions [12, 13]. Comparison of mortality from flu and COVID-19 is generally problematic due to the differences in data collection: in most countries deaths from COVID-19 must be recorded while deaths from influenza do not have to be recorded and are usually estimated from prevalence [14]. Research is currently going on to determine more accurate ways of estimating mortality from COVID-19 but it is believed to be much higher than for seasonal flu [13, 14].

Mathematical models can be used to inform and provide health decisions during a disease outbreak; besides they can be used to predict and perform peak detection of infected cases in a particular country [15]. A variety of models have been applied towards understanding the dynamics of COVID-19 [16].. These models can be broadly categorised into; Stochastic type models [17-19] and deterministic type [15, 20]. In the deterministic model, the population is usually divided into various compartments, namely Susceptible (S), infectious (I), Recovered (R) and Dead (D). Some models use all the compartments and are labelled SEIRD [21, 22]] while other models omit the Dead compartment and are SEIR [15, 23] or omit the exposed compartment and are SIRD [24 - 26] or end up with the basic SIR formulation by omitting the Exposed and Dead compartments [20, 27]. Irrespective of the number of compartments used above, models can be modified to include additional compartments like hospitalised, symptomatic, asymptomatic, among others [28 - 33]. In the context of deterministic models, are the so-called meta-population models which are capable of capturing the inherent heterogeneity of the populations, an aspect which cannot be done by compartmental models [16, 34]. In the early days of COVID-19 experts wondered if the disease would follow other virus pandemics and exhibit a second wave and possibly subsequent waves. Since then second waves have appeared in some countries on all continents; a few countries have even experienced third waves. This development has helped to promote mathematical modelling of the genesis and dynamics of second and subsequent COVID-19 waves [35 - 38]. In this paper we will use the SIRD model applied to a single wave.

In addition to mathematical models for the analysis of COVID-19 dynamics, there have emerged mathematical models that address intervention measures designed to control the rate of spread of the disease. As a starting point for these models, it is important to have an understanding of the baseline dynamics and hence estimates of the parameters associated with the unmitigated disease, since they are essential to our planning for initial interventions. Two of these parameters are particularly useful, namely, the transmission rate, *β*(*t*), which is the number of contacts per person per unit time, and the basic reproduction number, *R*_0_, already defined earlier. In most situations, intervention commences almost immediately COVID-19 emerges and the baseline parameters are estimated later, using data collected from the period preceding any major mitigation measures. The main purpose of the intervention is to reduce the contact rate, hence the reproduction number, so that the peak of infection reaches a level that can be managed by the available healthcare facilities and personnel. The mathematical models can broadly be classified into two categories. In the first category, variables and parameters associated with interventions are incorporated into the system of differential equations and hence they directly influence disease variables and parameters [28 – 29, 31 – 33, 39 - 40]. Although they are mathematically rigorous and elegant, they lead to more parameters that must be estimated; in addition, they tend to account for the effect of only a few intervention measures, whereas the impacts of interventions arise from all the measures taken together. In the second category, the intervention measures collectively, or a subset of them, are deemed to affect the transmission rate only, yielding expressions for the transmission rate which are piecewise continuous functions involving exponential, logistic, linear or constant functions [21, 26, 37, 42]. In this paper, we use the methods in the second category, since they are flexible and can easily be applied to develop scenarios, pending more rigorous investigation on the effect of the interventions.

We present the paper according to the following outline.

- In the next section we briefly describe the COVID-19 situation in Kenya and present the major NPIs and the timelines in which they were proposed.
- In Section 3, the SIRD model equations and initial conditions are given.
- In Section 4, we introduce a new method of estimating the parameters associated with the disease dynamics.
- A description is given in Section 5 of a mathematical model for interventions that takes into consideration the fact that interventions lead to a reduction of transmission rate, in the case of mitigation, or increase in the transmission rate, when some mitigation measures are lifted or when society violates prescribed mitigation regulations.
- In Section 6.1 we present solutions of the SIRD model, taking into consideration the mitigation and relaxation timelines.
- Section 6.2.1 presents the results of the effect of intervention measures of known magnitudes on the observed and predicted positivity rates.
- Using the observed positivity rates, in Section 6.2.2, we apply the method of planar envelopes around curves to deduce the model positivity trajectory, together with the mitigation and relaxation magnitudes that lead to the observed positivity rates.
- Finally, we give a few concluding remarks and recommendations in Section 7.

## 2 COVID-19 in Kenya

The rapidly spreading outbreak of the novel coronavirus in the African continent prompted the Kenya government to establish the National Emergency Response Committee on Coronavirus (NERC) on 28th February 2020 by Executive order. About 2 weeks later, the first case of COVID-19 in Kenya was confirmed on 13th March, 2020 [43] and has resulted in 100,773 confirmed infections and 1,763 deaths to 31st January 2021 [44]. Due to the rapid increase in cases, the Government of Kenya instituted several measures designed to curb the spread of the disease, while, at the same time, providing economic support to individuals identified as vulnerable. COVID-19 has hurt the Kenyan economy and the livelihood of the residents, despite the commendable steps taken by the government to alleviate the suffering of citizens. Concern has been raised, however, on the declaration by public and private insurers requiring COVID-19 patients to share in the cost of their diagnosis and treatment. Given that the costs are beyond the reach of many Kenyans, there have been reports of patients not seeking medical help at health centres for fear of being burdened with large bills, which they and their families would be unable to pay.

For our modelling, the strategies pursued can be divided into three periods, each with its distinct characteristics as outlined hereunder and also shown in Table 1.

**Table 1:** Strategies for mitigating COVID-19 in Kenya

|  | PERIOD 1<br>13 March to 8 April | PERIOD 2<br>9 April to 8 June | PERIOD 3<br>9 June to 8 August |
| --- | --- | --- | --- |
| CATEGORIES | ACTIONS | ACTIONS | ACTIONS |
| **1. CONGREGATIONS** |  |  |  |
| Bars and Clubs | Closed on 22 March | Closed | Closed |
| Places of worship | Closed on 22 March | Closed | Open from 6 July for 100 attendants per session |
| Funerals and Family gatherings | Limited numbers; social distancing and hygiene | Limited numbers; social distancing and hygiene | Limited numbers; social distancing and hygiene |
| Political and Social gatherings | Banned | Banned | Banned |
| Restaurants and Eateries | Open for limited hours for takeaway meals | Operate with social distancing and hygiene | Limited numbers; no alcohol from 31 July |
| Work from home | Compliance encouraged | Compliance encouraged | Compliance encouraged |
| **2. LEARNING INSTITUTIONS** |  |  |  |
| Schools | Closed | Closed | Closed |
| Tertiary Institutions | Closed | Closed | Closed |
| **3. RESTRICTION OF MOBILITY** |  |  |  |
| Cessation of Movement | None | For Nairobi, Mombasa, Kwale, Kilifi from 6 April | Lifted in 2 stages: 7 June and 7 July |
| Curfew | Country-wide overnight curfew from 27 March | Country-wide overnight curfew still in force | Country-wide overnight curfew still in force |
| Lockdown | None | For Eastleigh, Old Town Mombasa & Mandera | Lockdown lifted on 7 June |
| **4. PREVENTION** |  |  |  |
| Covid-19 Regulations | None | Published; criminal offence to contravene | Criminal offence to contravene |
| Public social distancing | Compliance encouraged | Compliance mandatory | Compliance mandatory |
| Public mask-wearing | Compliance encouraged | Compliance mandatory | Compliance mandatory |
| Public Hygiene | Compliance encouraged | Compliance mandatory | Compliance mandatory |
| **5. TRAVEL** |  |  |  |
| International air travel | A few allowed initially, later all suspended | Suspended | To resume on 1st August |
| Local air travel | Suspended 2 April | Suspended | Resumed on 7th July |
| Public transport (within the county) | To operate with social distancing & hygiene | To operate with social distancing & hygiene | To operate with social distancing & hygiene |
| Public transport (inter-county) | To operate with social distancing & hygiene | None to/from counties on cessation of movement | To operate with social distancing & hygiene |
| **6. ECONOMIC INCENTIVES** |  |  |  |
| Support for Vulnerable families | Plans to support the vulnerable | Money sent directly to vulnerable families | Money sent directly to vulnerable families |
| National Hygiene Programme (Kazi Mtaani) | Plans to support youth | Payment to youth for restoring public hygiene | Payment to youth for restoring public hygiene |
| General economic stimulus | Announced | Implemented | Implemented |

### Period 1 (13 March to 8 April)

Since COVID-19 was a novel disease, this period was spent in formulating policies and protocols on how to respond to it. A major decision was to immediately close learning institutions. Other mitigation measures were put in place, for instance: no overcrowding in public transport; social distancing and mask-wearing in public places; handwashing and sanitization at malls and supermarkets. There was low compliance and so a lot of time was spent appealing to residents to comply with the measures. In anticipation of the potential impact of the pandemic, the government introduced a tax break to provide some relief to residents. Despite the enacted mitigation measures, in most places it was business as usual. This period, therefore, can safely be regarded as the time when covid-19 was still unmitigated. Hence our model treats this period as the baseline, since no significant impact of mitigation measures had been realised.

### Period 2 (9 April to 8 June)

Due to the rapid increase in cases, and the public’s relaxed attitude to COVID-19, the government took a move to enforce the mitigation measures by enacting COVID-19 regulations whose contravention was a criminal offence [45]. It also published proto-cols to govern the operations of restaurants and eateries. To provide an economic incentive to a category of residents the government implemented two programmes to help the vulnerable and youth. The period can be regarded as one of the application of mitigation measures, despite attempts by a cross-section of society to flout the rules. Consequently, our model treats this period as that of mitigation.

### Period 3 (9 June to 8 August)

Enforcement of the mitigation measures during Period 2 had a devastating effect on the country’s economy and people’s livelihoods. Many industries and small businesses laid off workers or simply folded. Other establishments placed workers on half salary or gave leave without pay while waiting for the situation to stabilize. To ease the hardship being experienced, the government gradually relaxed some of the mitigation measures. There was discussion about opening learning institutions in September but the idea was shelved on based on the trend of the pandemic. Our model treats this period as that of gradual relaxation of control measures.

Table 1 gives a summary of the major actions taken during each of the three periods. The information in this table was obtained from the Ministry of Health, Kenya [44] and Presidential Addresses on COVID-19 [46]. Most of the information was also available from Academia Kenya [47] and in the dailies.

Figure 1 shows the trend of the 7-day moving averages of numbers and percentages of three para-meters, that is, infections, deaths and recoveries, from 13th March 2020 for numbers and 25th March 2020 for percentages, to 31st January 2021. The percentages were of the numbers of daily observed variables relative to the daily number of people tested on that day. It is argued, with some justification that more realistic percentages can be obtained by computing the percentages relative to test numbers lagged by a week or two, since people do not necessarily get infected, recover or die on the same day they are tested.

**Figure 1:**
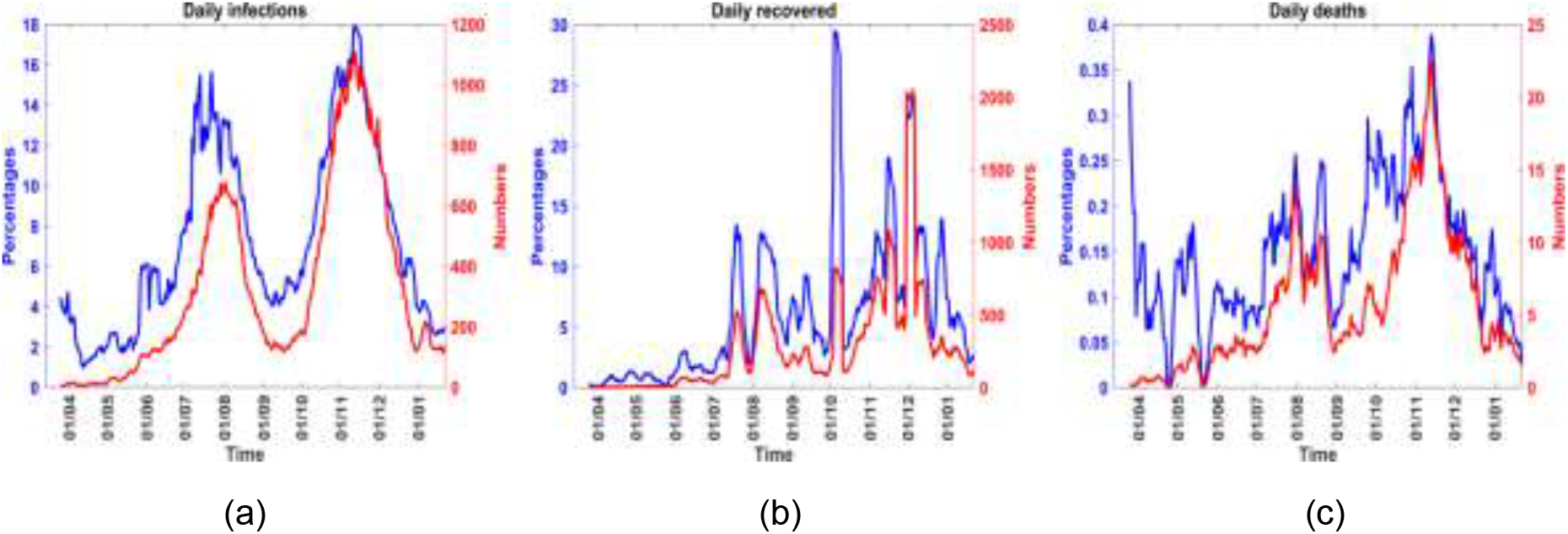
The 7-day moving averages of numbers and percentages of three variables, from 13_th_ March 2020 for numbers and 25_th_ March 2020 for percentages, to 31st January 2021.

In Figure 1(a), the infections increase gradually and reach a maximum in mid-July, with a positivity rate of just under 16%. The infections then decrease until early September when they begin to increase thus indicating the onset of the second wave, which peaked in early November with a positivity rate of about 18% and prompted fresh mitigation measures to be put in place from 4th November 2020. Figure 1(b) shows that recoveries also follow the wave pattern of the infections but there exist considerable fluctuations, with significant spikes, possibly due to accumulated data not accounted for in previous days. The fluctuations and spikes can also be partly attributed to uncertainty in obtaining data on recoveries from home-based care which was in effect from July 2020. Figure 1(c) shows that the deaths also exhibit the wave pattern of the infections but they fluctuate considerably but are generally on the low side, with the 7-day averages not exceeding 25 in numbers or in percentages.

## 3 SIRD model formulation

In this article, we consider a SIRD mathematical model. We assume a homogeneous mixing in the population. At the time, *t*, the population is divided into four classes; Susceptible, infectious, recovered and the dead, denoted respectively by, *S*(*t*), *I*(*t*), *R*(*t*) and *D*(*t*), as shown in Figure 2 Since this is a new disease, there is no prior immunity, hence everybody is susceptible to COVID-19. Upon being infected with the disease, susceptible individuals move to the infectious class, from which they either recover or die from the infection.

**Figure 2:**
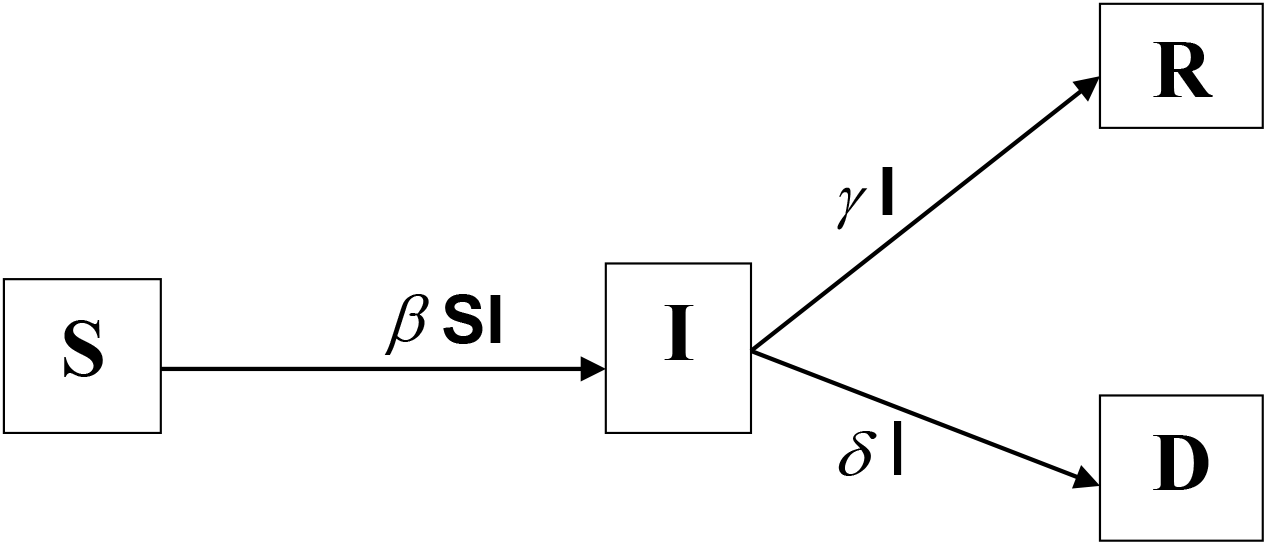
Compartmental SIRD model.

We assume that the total population, *N* is constant over time. For simplicity we assume that the variables are already normalised on division by *N* such that

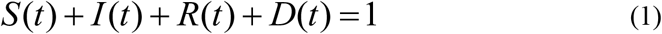

In the presentation of our results, the variables will be given as convenient in terms of actual observed numbers or proportions or percentages or as fractions of the total population.

The mathematical equations describing the movement of individuals in different compartments are given by:

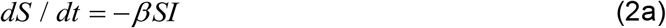

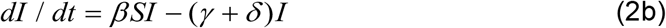

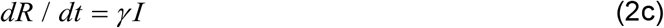

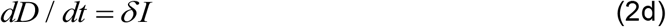

The system in Equation (2) is solved subject to the initial conditions: *S*(0) = *S*_0_, *I*(0) = *I*_0_, *R*(0) = *R*_0_ and *D*(0) = *D*_0_, where S_0_, I_0_, R_0_ and D_0_, are the initial proportions of the Susceptible, Infectious, Recovered and Dead, respectively. At the very start of the epidemic, there is one infected individual so that I_0_ = 1/N. At this stage there are no recoveries or deaths so that R_0_ = D_0_ = 0 and Equation (1) yields S_0_ = 1-1/N. The SIRD model has been applied to seasonal influenza [48 - 50] and COVID-19 [20, 24 – 26], among other publications.

## 4 Parameter estimation

To solve the system in Equation (2) subject to appropriate initial values, it is necessary to compute the parameters *γ, β* and *δ*. This is usually by use of optimization software [51 – 53], preceded by specifying approximate values for the parameters, or the interval in which the parameter values lie, or by use of Monte-Carlo methods to select initial values of the parameters at random and facilitate computation of the optimum values using methods based on diverse mathematical concepts. The process sometimes is long and may require hundreds, if not thousands, of iterations. In this paper, we propose a new approach to determining initial estimates of the parameters.

Observational or experimental values are usually available at discrete points in time denoted, *t*_0_, *t*_1_, *t*_2_, …, *t*_max_, where *t*_max_ is the maximum time for which the disease data is available. Starting from the initial values, *S*(0) = *S*_0_, *I*(0) = *I*_0_, *R*(0) = *R*_0_ and *D*(0) = *D*_0_, we find that at the current iteration, *t*_*k*_, Equation (1) yields

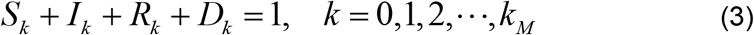

where *S*_*k*_ = *S* (*t*_*k*_), *I* _*k*_ = *I* (*t*_*k*_), *R*_*k*_ = *R*(*t*_*k*_) *and D*_*k*_ = *D*(*t*_*k*_) and *k*_M_ is the maximum number of time steps or nodes.

Given the significance of infection in understanding the dynamics of the disease, data on the infections, *I*_*k*_, is often available to a considerable degree of accuracy and reliability. Whereas data on the deceased, *D*_*k*_, and the recovered, R_k_, may be available, it is often less reliable, particularly in countries with limited health facilities. In situations where only data on the infections is given, it is possible to estimate the recovered and deceased by integrating Equations (2c) and (2d) to yield:

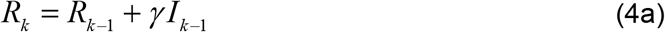

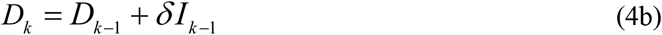

where *γ* and *δ* are the recovery and death rates, respectively, and are to be estimated.

From Equation (4a) using k = 1 yields *R*_1_ = *γ I*_0_, since *R*_0_ = 0. Hence *R*_1_ is dependent on the observed value *I*_0_. Using k = 2, we conclude that *R*_2_ is dependent on the observed values *I*_0_ *and I*_1_. We can show that in general, *R*_*k*_ is dependent on the observed values *I*_0_, *I*_1_, *I*_2_, …, *I*_*k* −1_. Analogously, we can show that *D*_*k*_ is dependent on the observed values *I*_0_, *I*_1_, *I*_2_, …, *I*_*k* −1_. Although the quantities *R*_*k*_ and *D*_*k*_ are not observed values, they are dependent on observed infections and, to meet our computational objectives, they are regarded as “observed recovered fraction” and “observed dead fraction”, respectively. Similarly, values of the susceptible fractions, *S*_*k*_, are rarely availed, except in a highly controlled environment, for instance, a school setting [54]. Despite this, by solving for *S*_*k*_ in Equation (3), we obtain a value of the susceptible which is dependent on observed infections and will also be regarded as “observed susceptible fraction”.

To illustrate our approach for estimating the parameters, we assume that the deceased values, *D*_*k*_ are given and the death rate, *δ*, is approximated by the Case Fatality Rate (CFR) or any other suitable method. Consequently, we consider Equations (2) and (4a) only and estimate *γ* and *β*. Every disease has associated with it a number of days of recovery, i.e. the days for which the patient remains contagious after being diagnosed with the disease; the inverse of those days is the recovery rate, *γ*. For instance, the recovery days from influenza are on average 5 – 7 days [12 – 13]; hence *γ* for influenza is on average 1/7 to 1/5. The recovery days from mild to moderate COVID-19 are on average 10-14 days [12 - 13]; hence *γ* for this type of COVID-19 is on average 1/14 to 1/10. To begin our computation, we assume that the recovery days for COVID-19 do not exceed d_M_ days, where d_M_ is an integer that is large enough to preclude any medical evidence that a patient of COVID-19 could still be contagious beyond d_M_ days, for instance 100. Hence the recovery rate, *γ*, will be in the interval (1/d_M_, 1). We now establish grid point, or nodes, that *γ*_*i*_, in the interval (*d*_*M*_, 1) such that

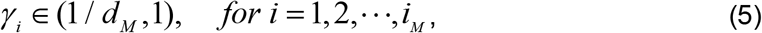

where *i*_*M*_ is the maximum number of node pertaining to *γ*.

At the time step *k*, insert *γ*_*i*_ in Equation (4a) and then Equation (4a) into Equation (3) to find

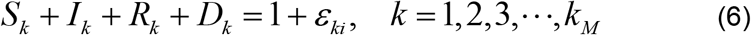

where *ϵ*_*ki*_ is the error, at the grid point *t*_*k*_, associated with observation and also with the value *γ*_*i*_ If *ϵ*_*ki*_ is close to zero then *γ*_*i*_ is close to the recovery rate for the disease.

The transmission rate, *β*(*t*), for COVID-19 is a finite non-negative number, and must therefore be such that 0 *< β*(*t*) *< β*_M_, where *β*_M_ is a positive number and is chosen large enough to include the highest possible transmission rate of COVID-19. We then establish grid points, or nodes, *β*_*j*_, in the interval (0, *β*_M_) such that

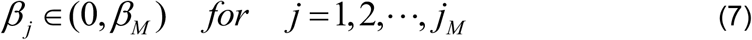

where *j*_*M*_ is the maximum number of nodes pertaining to *β*.

We now use *γ*_*i*_ from Equation (5) and *β*_*j*_ from Equation (7) together with the value of *δ* estimated from CFR, to solve the system (2), at the time step *k*. This will yield values which we denote *S*^*c*^, *I*^*c*^, *R*^*c*^ and *D*^*c*^, where the superscript signifies values from computation to distinguish them from observed *S*_*k*_, *I*_*k*_, *R*_*k*_ *and D*_*k*_. Analogous to Equation (6), the values from computation satisfy

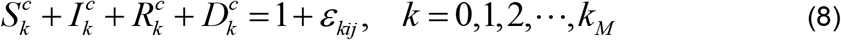

where *ϵ*_*kij*_ is the error, at the grid point *t*_*k*_, associated with observation together with the values *γ*_*i*_ and *β*_*j*_. If *ϵ*_*kij*_ is close to zero then *γ*_*i*_ and *β*_*j*_ are close to the recovery and transmission rates, respectively.

Subtracting Equation (6) from (8) yields

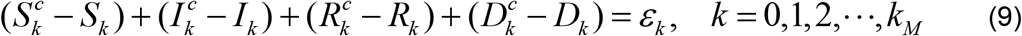

where *ϵ*_*k*_ is the total error at the time step *t*_*k*_. We note that each of the terms in brackets on the LHS is the error in the corresponding variable.

The disease variables are nonlinear functions of the parameters *γ*_*i*_ and *β*_*j*_, hence it is not advisable to compute the correlation coefficients between the computed (predicted) and the observed variables to determine how closely the computed and observed values agree. Hence we use an error metric based on the time average of the individual errors and we choose to minimize the error metric. There are a number of possible error metrics but we choose to use the Root Mean Square Error (RMSE), although we could also have used the Mean Absolute error (MAE) [55]. For RMSE we determine *γ*_*i*_ *and β*_*j*_ to obtain

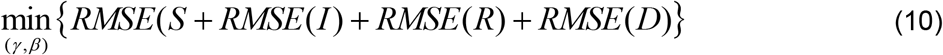

where for the variable X,

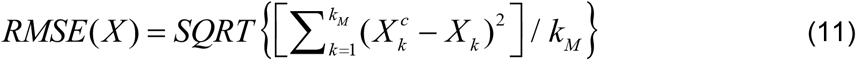

Starting from initial values of *γ*_*i*_ and *β*_*j*_ it is possible to institute search procedures that will lead to the minimum in Equation (10), for instance [51 – 53]. The amount of effort in reaching this minimum will depend on how close the initial values are to the actual solutions. To narrow down the intervals of the search, we use a different approach based on the fact that the disease parameters, as used in the formulations leading to Equation (2), are spatial and temporal averages. We begin by determining the maximum sum inside the curly brackets in Equation (10). We then express the sum at each point as a percentage of this maximum sum; consequently, the minimum values will be associated with the least percentages. Thereafter, we average the disease parameters within regions bounded by concentric circles whose radii are the percentages. The regions are expanded from the innermost circle, associated with the least percentage, to larger circles associated with higher percentages, while keeping track of the sum in Equation (10). Initially this sum is large but decreases as we expand the percentage of averaging; eventually the sum reaches a minimum and begins to increase again as the percentage of averaging continues to increase. The minimum in Equation (10) is at values of the parameters associated with the turning point. To ensure that we identify all the minima, it is important to start from the least percentage and proceed till 100%, or close to it. This way it will be possible to identify which minima are local and which one is global. The best that can be done with this averaging technique is to obtain intervals within which lie isolated values of *γ*_*i*_ and *β*_*j*_ that lead to the suspected minimum of the error matrices in Equation (10). Thereafter, other search and optimization procedures can be applied to obtain more accurate parameter values, if need be [51 – 53]. Here again, we propose a slightly different approach: simply divide the interval so identified into smaller subintervals to establish a denser network of grid points. We then go through the computations leading to the sum in Equation (10) for all paired values of *γ* _*i*_ and *β* _*j*_. This time, we use a procedure that searches for a minimum value from an array and *j* identifies the associated *γ* _*i*_ and *β* _*j*_. The new subinterval, within which lie the appropriate *γ*_*i*_ and *β* _*j*_ can again be divided into smaller subintervals and the process repeated until required accuracy in *γ* _*i*_ and *β* _*j*_ is obtained.

## 5 Modelling intervention

In this paper we formulate an intervention model which leads to piecewise exponential functions for the transmission rate. We assume that the recovery rate, *γ*, and the death rate, *δ*, do not change during the interval of the intervention. Our model takes into account the fact that intervention not only leads to a reduction of the transmission rate, through mitigation, but can lead to a surge in the transmission rate, through relaxation of, or non-compliance with, mitigation measures.

Let the daily events be at the time nodes denoted *t*_0_, *t*_1_, *t*_2_, …. Suppose intervention is initiated at the time node *t*_*k*_ then there will be a difference in the transmission rate before and after *t*_*k*_. Let *β_b_*(*t*) be the transmission rate before, and up to, the time *t*_*k*_; the quantity *β*_*b*_(*t*) could be the result of baseline dynamics or it could be due to the dynamics from some immediately preceding intervention. Associated with *β*_*b*_(*t*) is a reproduction number which we label R_b_(t). If this R_b_(t) is due to baseline dynamics then it is the basic reproduction number R_0_ ; otherwise it is an effective number, R_e_,

We now assume that for any time *t > t*_*k*_, the rate of decrease of the transmission rate, as a result of intervention measures, is proportional to the transmission rate at that time. This yields the following general solution for the effective transmission rate;

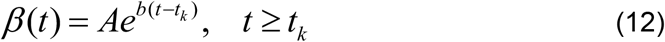

The main objective is to gradually change the transmission rate at the time of intervention, namely, *β*_*b*_(*t*_*k*_), by a fraction *c* so that the effective transmission rate at a future time, say *t*_*k*+*m*_, where *m >* 0, becomes (1 *− c*)*β*_*b*_(*t*_*k*_). To determine the constants *A* and *b* in Equation (12), we impose the conditions

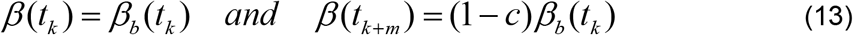

We need to consider what happens after the objective of the intervention has been met, namely for *t > t*_*k*+*m*_. It is reasonable to assume that after the objective of a particular intervention has been met, the transmission rate will remain constant at the level already achieved, until another intervention takes place. The choice of *m* which enables this to be achieved will depend on the implementation goals. When an intervention takes place on day *t*_*k*_, the optimum transformation of the transmission rate, hence of the reproduction number also, does not occur instantly but takes place say *m* days later, that is, at the time *t*_*k*+*m*_, where *m >* 0. In fact it is a common observation that following an event that spreads COVID-19, an increase in infections is usually observed about 7 to 14 days later. It is, therefore reasonable to select *m* in, or close to, the interval 7 to 14 days. Using Equation (13) and the explanation for what happens beyond *t*_*k*+*m*_, we obtain the following expression for the transmission rate, before and after the intervention.

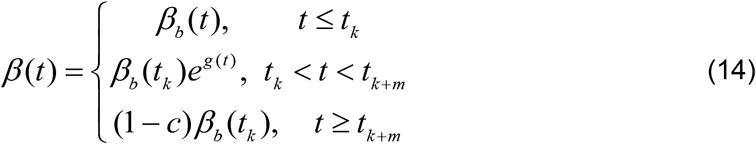

where

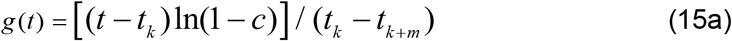

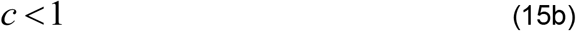

The last equation in Equation (14) gives the optimum value of the effective transmission rate that will be achieved due to intervention. At any given time, t, the effective reproduction number, *R*_*e*_, is computed from

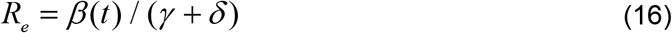

From the last equation in Equation (14) we note that when 0 *< c <* 1, then *β*(*t*_*k*+*m*_) *< β*_*b*_(*t*_*k*_) ; this corresponds to the intervention being a mitigation, since it yields a smaller future transmission rate which represents a reduction by a fraction *c* of the transmission rate at the time of intervention before mitigation occurred; we call the quantity 100c the “percent mitigation”. On the other hand when *c <* 0 then *β*(*t*_*k*+*m*_) *> β*_*b*_(*t*_*k*_); this corresponds to the intervention being a relaxation of the existing mitigation measures since it yields a larger future transmission rate which represents an increase by a fraction |*c*| of the transmission rate at the time of intervention; we call 100 × |*c*| the “percent relaxation”. In computing *R*_*e*_ the values of *γ* and *δ* remain fixed and only the value of *β* changes. Consequently the percentage changes in *β*, as indicated above, also apply to *R*_*e*_. Previous researchers restricted c to the interval [0, 1]; by Equation (15b), we extend c to negative values to account for spikes in the dynamics that may occur as a consequence of relaxing mitigation measures.

The parameter *c* is important for this type of intervention modelling and yet only two of its values are obvious, namely, *c* = 0 implies the absence of any control and *c* = 1 is the unlikely scenario of absolute control where there is no disease transmission. The other values of *c* are more complicated to determine. The most thorough method is to identify all the interventions that impact on the disease transmission, hence contribute to *c*, and assign weights to their impacts. The parameter *c* can then be computed as the weighted average of the impacts. Groups of the impacts could then be isolated to obtain their relative contribution to the decrease or increase in the disease transmission. Table 1 lists some of the intervention measures and related effects that could be taken into account in this exercise: mask-wearing, closure of learning institutions, curfews, travel restrictions, limitations on gatherings, restrictions on operations of bars and restaurants, economic activities, social distancing, availability of PPEs and hospital space etc. To assess the impact of all these factors on disease transmission requires a truly collaborative effort involving a multidisciplinary team. Once the value of *c* is estimated, it can be described in simple terms for public health implementation. For instance, two intervention scenarios are mentioned in [23], namely: *moderate lockdown*, regarded as the intervention which reduces transmission by 25% during lockdown followed by transmission at 90% of the pre-lockdown value; and *hard lockdown*, regarded as the intervention which reduces transmission by 44% during lockdown followed by transmission at 90% of the pre-lockdown value. In terms of our formulation, *moderate lockdown* is equivalent to mitigation with *c* = 0.25 followed by relaxation with *c* = *−*2.6 while *hard lockdown* is equivalent to mitigation with c = 0.44 followed by relaxation with *c* = *−*1.05.

## 6 Results

The models were developed and computations carried out, per timelines associated with government and public response to COVID-19, as specified in Section 2. We first present results for solutions of the SIRD system using the new method described in Section 4; then we present results for the intervention model described in Section 5. COVID-19 data was obtained from the following sources: Worldometer [1], Ministry of Health, Kenya [44], World Health Organization [56], and Our World in Data [57].

### 6.1 Solution of SIRD system across different intervention periods

In this section we solve the system of equations using the methods described in Section 4 for the periods identified in Section 2: Period 1 (Baseline), Period 2 (Mitigation) and Period 3 (Relaxation). The objective is to determine whether there are any differences in the disease parameters among the three periods. In Table 2 we list the values of parameters and initial conditions used during various periods of computation.

**Table 2:** Parameters and initial conditions at baseline, mitigation and relaxation periods

| Quantity | Symbol | Baseline Value | Mitigation Value | Relaxation Value |
| --- | --- | --- | --- | --- |
| Population | N | 5586 | 100683 | 353727 |
| Initial Infections | I(0) | 0.1790190E-04 | 0.9932163E-05 | 0.2827039E-05 |
| Initial Recovered | R(0) | 0 | 0 | 0 |
| Initial Deaths | D(0) | 0 | 0 | 0 |
| Initial Susceptible | S(0) | 0.999982098 | 0.999990067 | 0.999997173 |
| Number of days | ----- | 27 | 88 | 149 |
| Maximum time nodes | $k_M$ | 27 | 88 | 149 |
| Maximum days of recovery | $d_M$ | 100 | 100 | 100 |
| Least value of $\gamma$ ($=1/d_M$) | $\gamma_M$ | 0.01 | 0.01 | 0.01 |
| Subintervals for $\gamma$ | ----- | 1000 | 1000 | 1000 |
| No. of nodes for $\gamma$ | $i_M$ | 1000 | 1000 | 1000 |
| Largest value of $\beta$ | $\beta_M$ | 1 | 1 | 1 |
| Subintervals for $\beta$ | ----- | 750 | 750 | 750 |
| No. of nodes for $\beta$ | $j_M$ | 750 | 750 | 750 |

For all the periods, we assumed that covid-19 patients are unlikely to be contagious after 100 days, so that d_M_ = 100, hence *γ* ∈ (0.01,1). We also assumed that *β* ∈ (0,1) for covid-19, since the choice of a larger upper limit would not make any difference to the results. To obtain the initial estimates of *γ* and *β*, by the averaging method described in the last paragraph of Section 5, the intervals (0.01, 1), for, *γ* was divided into 1000 subintervals while the interval and (0, 1), for, *β* was divided into 750 subintervals, and a node located in the middle of each subinterval. On identification of the appropriate initial estimates, the subintervals in which they lay were each further divided into 400 subintervals, to establish 400 nodes, and the MIN function in MATLAB was used to solve Equation (10). Further subdivision was not required, since the values obtained agreed to 5 significant figures.

#### (a) Baseline Solutions

For results involving the baseline, computation is carried out from 13th March to 8th April, 2020, with projections made till stability is reached. Use is made of the parameters and initial conditions given in the baseline column of Table 2. Computed values of *γ* and *β*, are given, to 5 significant figures, in the baseline row of Table 3. Also given are related parameters and results. The basic reproduction number is 2.76 which is consistent with results from other computations, e.g. [23].

**Table 3:** Parameters and related quantities for different disease periods. Recovery rate is denoted *γ,δ* denoted case-fatality proportions(CFR), *β*, transmission rate and 1*/γ* denotes recovery days, R=*R*_0_, basic reproduction number (for Baseline) and R=*R*_*e*,_ effective reproduction number (for mitigation and relaxation)

| | $\gamma$ | CFR ($\delta$) | $\beta$ | $R$ | $1/\gamma$ | Peak infection | RMSE sum |
| --- | --- | --- | --- | --- | --- | --- | --- |
| Baseline | 0.051750 | 0.015 | 0.18448 | 2.7638 | 19.3 | 0.270 | 0.00255 |
| Mitigation | 0.055294 | 0.0344 | 0.14852 | 1.6558 | 18.0 | 0.0916 | 0.000570 |
| Relaxation | 0.015099 | 0.0297 | 0.092675 | 2.0687 | 66.2 | 0.1655 | 0.000804 |

Figure 3(a) shows the observed and computed cumulative infection numbers and percentages during the baseline period; the agreement is quite good. Although results are available for the parameters S, I, R and D, we present results only for I, since it is the most important variable we use in our subsequent analyses.

**Figure 3:**
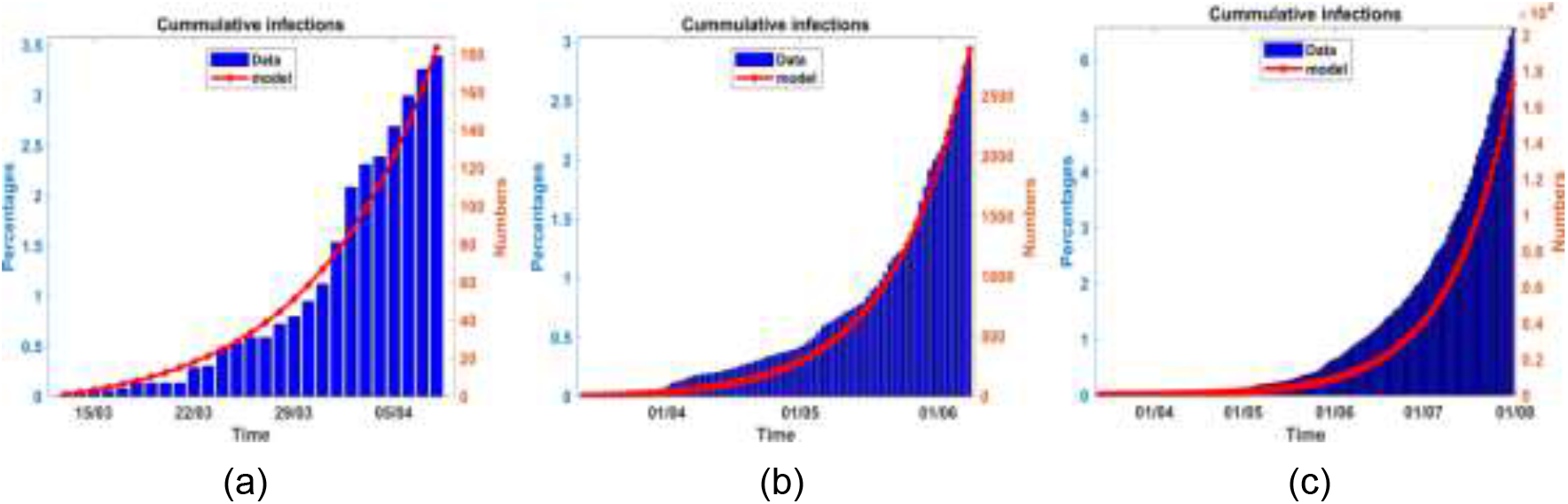
Observed and computed cumulative infected proportions and numbers at baseline (3(a)), observed and computed cumulative infected proportions and numbers up to mitigation period (3(b)) and Observed and computed cumulative infected proportions and numbers up to relaxation period (3(c)).

Figure 4 shows the graphs for infection percentages obtained using data up to the end of the baseline, mitigation and relaxation periods. The graph labelled baseline dynamics shows the infection percentages associated with the baseline data. It achieves a peak of approximately 27% in late May 2020, thus indicating the way the dynamics would have proceeded if mitigation measures were not put in place on 8th April 2020.

**Figure 4:**
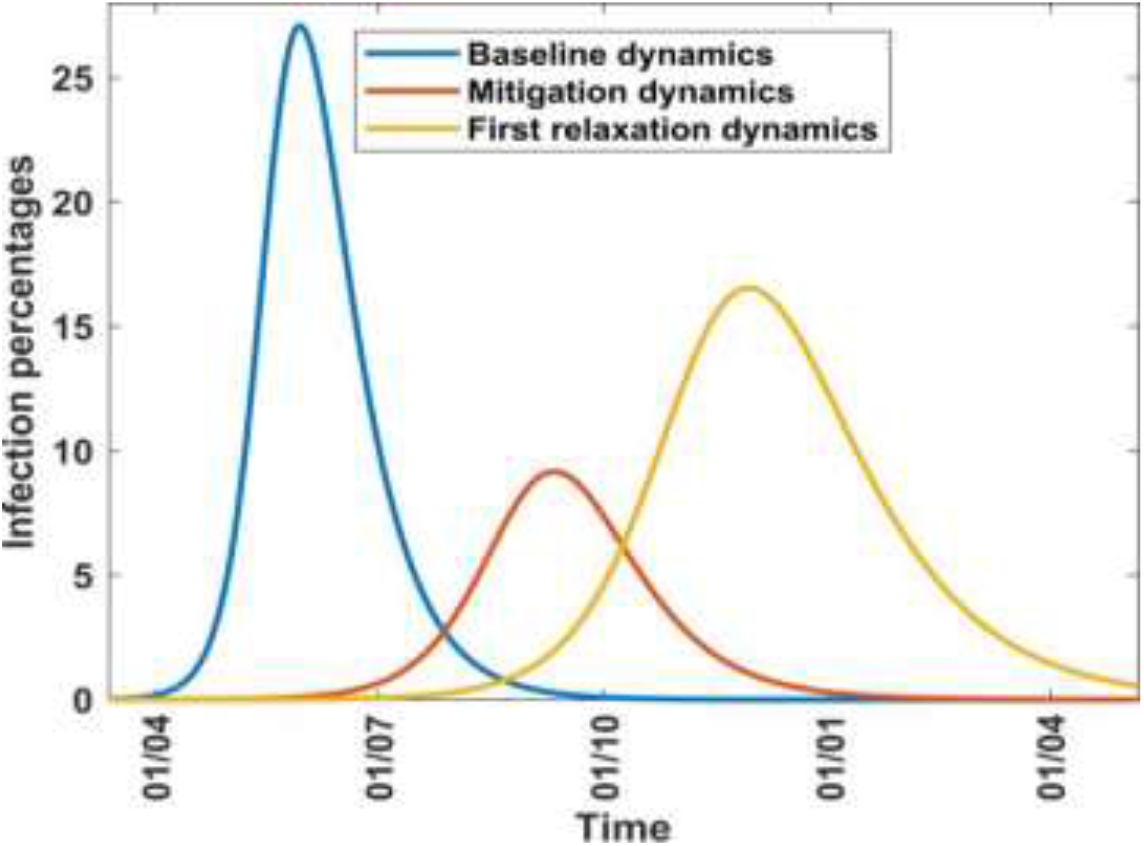
Infection proportions during baseline, mitigation and relaxation periods.

#### (b) Mitigation Solutions

For results involving mitigation, computation is carried out from 13th March to 8th June. The objective of the computation is to find out whether the inclusion of data during the mitigation period made any difference in the rate of transmission of the disease and the dynamics of the infection. Use is made of the parameters and initial conditions given in the mitigation column of Table 2. Computed values of *γ* and *β*, are given, to 5 significant figures, in the mitigation row of Table 3. Also given are related parameters and results. The results indicate that the reproduction number was reduced by about 40% from 2.76 at the baseline to 1.66 at mitigation. These results indicate that the mitigation measures that were put in place on 8th April helped to reduce the rate of spread of the disease. Other assessments showed, however, that the gains in controlling the disease were met at a considerable economic and social impact on the country. In Figure 4 the graph labelled mitigation dynamics shows the infection percentages associated with data up to the end of the mitigation period. This graph peaks at 9.2% thus indicating that the mitigation measures put in place from 9th April to 8th June had reduced the peak infection to a level more manageable for healthcare. Figure (3b) shows the observed and computed cumulative infection numbers and percentages during the mitigation period; the agreement is quite good.

#### (c) Relaxation Solutions

For results involving the relaxation, computation is carried out from 13th March to 8th August. The objective of the computation is to find out whether the inclusion of data during the relaxation period made any difference in the rate of transmission of the disease and the infection dynamics. Use is made of the parameters and initial conditions given in the relaxation column of Table 2. Computed values of *γ* and *β*, are given, to 5 significant figures, in the relaxation row of Table 3. Also given are related parameters and results. The results indicate that the reproduction number increased by about 25% from 1.66 at mitigation to 2.07 at relaxation, thus showing that relaxation of mitigation measures led to an increased rate of transmission of the disease. Figure 3(c) shows the observed and computed cumulative infection numbers and proportions during the relaxation period; the agreement is good but not as well as in the previous two cases. In Figure 4 the graph labelled relaxation shows the infection percentages, associated with the relaxation period, which peak at about 16.6%. The implication is that the lifting of mitigation measures on 8th June, increased the peak infection to a level that could put more pressure on healthcare facilities, although not as much as would have happened with unmitigated disease. These results indicate that lifting the mitigation measures on June 8th subsequently placed an increased disease burden on society despite the temporary relief from the adverse economic and social effects due to mitigation. Figure 4 shows that if mitigation measures were not lifted, the disease would virtually disappear by January 2021 but relaxation shifts the disease wave and makes it disappear in mid-2021.

### 6.2 Results from modelling interventions

In the previous subsection, we presented results based on the solution of the SIRD system of equations, in which interventions had taken place and we aimed to determine how much they affected the transmission rate, reproduction number, infection rates and other variables. In this subsection, we would like to solve the following two problems:

- Given the magnitudes of the mitigation and relaxation measures, determine their effects on the disease dynamics, especially the positivity rates.
- Given the positivity rate curve, determine the magnitudes of the mitigation and relaxation measures that could have resulted in the curve.

A solution to both problems requires the application of the methods described in Section 5. It necessitates setting up scenarios hence the results are largely qualitative and yield only broad outcomes for use in the initial planning and further investigations. We make the following assumptions:

i. The largest portion of intervention measures is put in place at the beginning of the intervention period, namely close to 8th April 2020 for mitigation and 8th June 2020 for relaxation. Inevitably, there will subsequently be minor adjustments to the interventions which, if they were significant enough, would be regarded as independent interventions in their own right.
ii. The effect of the intervention, resulting in the optimum values of the transmission rate and effective reproduction number, is not achieved immediately but takes some days, normally up to 21 days but mostly between 7 to 14 days.
iii. Once the optimum values of the transmission rate and effective reproduction number are achieved, they govern the disease dynamics till another intervention takes place, either a mitigation or a relaxation.

#### 6.2.1 Determination of positivity rates given intervention magnitudes

A solution to the problem here requires information on the magnitudes of the intervention measures, hence appropriate values of the parameter c to be used in Equation (14). In case there is a good accounting of the factors that contribute to the disease transmission rate, and their relative effects, we can readily determine the parameter *c* as indicated in Section 5. If necessary, the parameter c can be determined by solving the SIRD model as done in Section 6.1. Ideally, this problem should be solved *a priori*, namely before any interventions are put in place, so that the outcomes can be used to plan for the interventions. After interventions have occurred, the problem can be solved to learn what action ought to have been taken.

##### (a) Results from modelling mitigation

As pointed out earlier, the first incident of the disease in Kenya was on 13th March 2020. If the disease was left unmitigated it would have spread as indicated in the blue baseline dynamics curve in Figure 4. The infection would have peaked in late May 2020 and virtually disappeared by early October 2020. The associated parameters and quantities are indicated in the baseline row of Table 3. These values indicate that slightly over a quarter of the population would have been infected at the peak of the disease, a fact which would have placed a considerable burden on health facilities. Mitigation was consequently effected on 8th April 2020 and had the effect of slowing down the spread of the disease.

To study the effect of different mitigation strategies during the period 8th April to 8th June 2020, we can develop scenarios by varying the parameter c in Equation (15). Since the mitigation is on the baseline dynamics, the transmission rate up to the time of the mitigation, *β*_*b*_(*t*), must be the baseline transmission given in the baseline row of Table 3, namely, *β*_*b*_(*t*) = 0.1845. Using this value of *β*_*b*_(*t*) in Equation (14), with m = 15, and drawing the curves associated with different values of c, we obtain Figure 5. For a given value of c the curve shows the trajectory of the infection as a result of a one-time mitigation force on 8th April 2020, with no other subsequent interventions. It can be seen that the infection peaks reduce as the value of c increases and they tend to occur later, meaning that the more stringent the mitigation measures, the lower will be the peak infections but they will occur at a later time.

**Figure 5:**
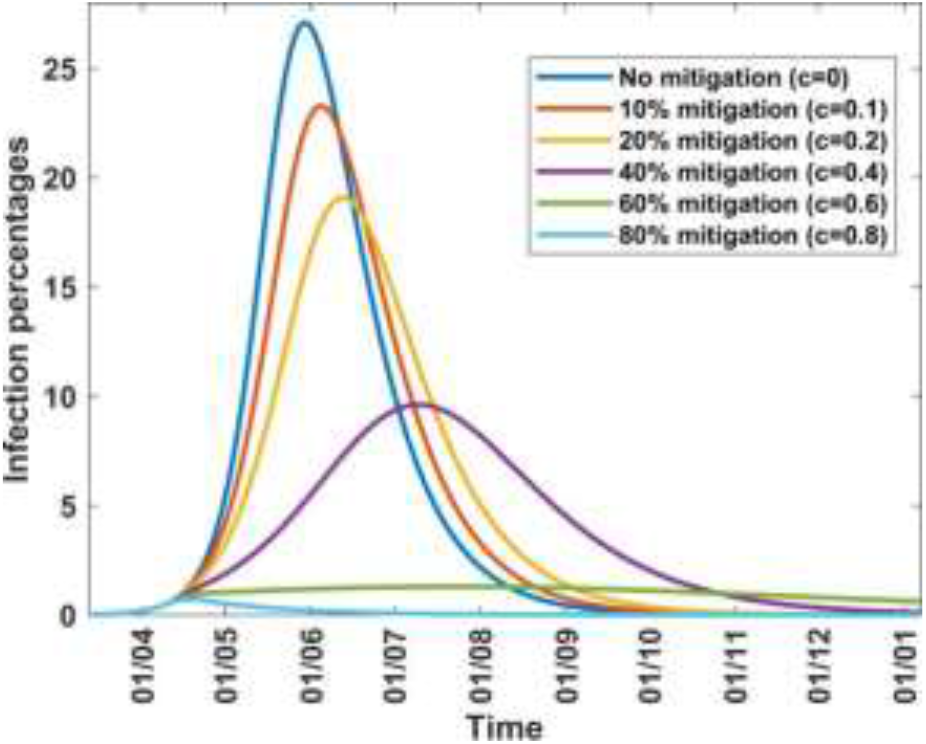
Infection percentages for various mitigation scenarios.

Table 4 shows how varying the parameter c affects the transmission rate, the effective reproduction number, from Equation (16), and the infection. It is noted that as *c* increases, the peak infection and the effective reproduction number decrease. Further increase shows that for *c* = 0.8, the peak infection is 1.2% and *R*_*e*_ = 0.55 *<* 1, meaning that there is no disease spread. This situation represents taking mitigation action which is so drastic that the disease is suppressed; the consequences to society for such action can be grave and so disease suppression is not a practical option. Table 4 also gives the infection at the end of the mitigation period, namely 8th June 2020. It enables the planner, who on 8th April 2020 is proposing a 2-month mitigation action, to forecast the infection percent on 8th June 2020, depending on the force of the mitigation.

**Table 4:**
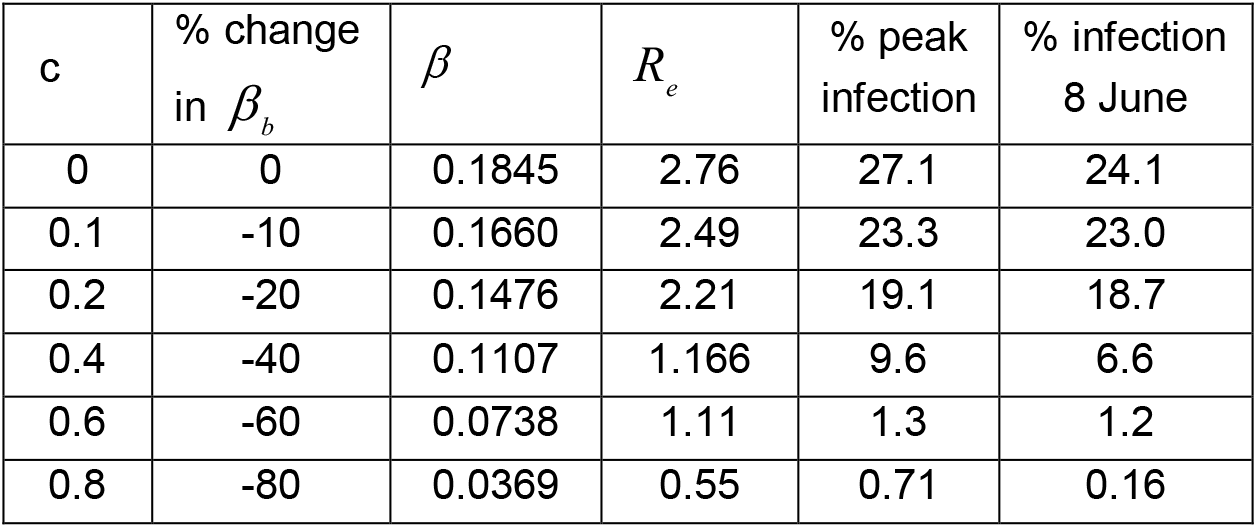
Scenarios involving changes in the parameter c under mitigation from baseline. The following values are used *β*_*b*_ = 0.1845, *R*_*b*_ = 2.76, *γ* = 0.0518 and *δ* = 0.01

A question of interest is: Can we quantify the impact of the mitigation measures effected from 8th April to 8th June? To answer this question *a priori* we would have to estimate, before 8th April 2020, the value of the parameter c as described in Section 5. We can also answer this question *a posteriori*, namely, after 8th June 2020, by making use of the results in Section 6.1 where we solved the system of equations from the onset of the disease up to the end of the mitigation, namely, from 13th Mach 2020 to 8th June 2020. Table 2 shows that from the baseline period to the mitigation period the basic reproduction number decreased from 2.76 to the effective reproduction number of 1.66. This change represents a decrease of approximately 40% between the two periods. It is logical, therefore, to take *c* = 0.4 as the mitigation parameter in Equation (14) and the trajectory of the infection would be the curve identified by 40% mitigation in Figure 5.

##### (b) Results from modelling relaxation

As a result of the adverse effects of mitigation, the government decided to relax some of the mitigation measures from 8th June to 8th August 2020. This is the period we regard as the “relaxation period” in our model; it is characterised by an increase in the transmission rate, reproduction number, and infection. If we have a good accounting of the relaxation factors that contribute to the disease transmission rate and can estimate their relative effects, we can determine the parameter c to be used in Equation (14), as described in Section 5; the values of c will be negative, since they imply an increase in transmission rate above reference values. The scenarios cannot, however, be based on the baseline dynamics; they must start from an appropriate mitigation scenario which ended on 8th June 2020. We have established in the previous paragraph that such a scenario corresponds to mitigation with c = 0.4. Consequently, to study the effect of different relaxation strategies from 9th June to 8th August 2020, we develop scenarios by varying the parameter c, taking into consideration the fact that the scenarios are dependent upon 40% mitigation, as shown in Figure 6.

**Figure 6:**
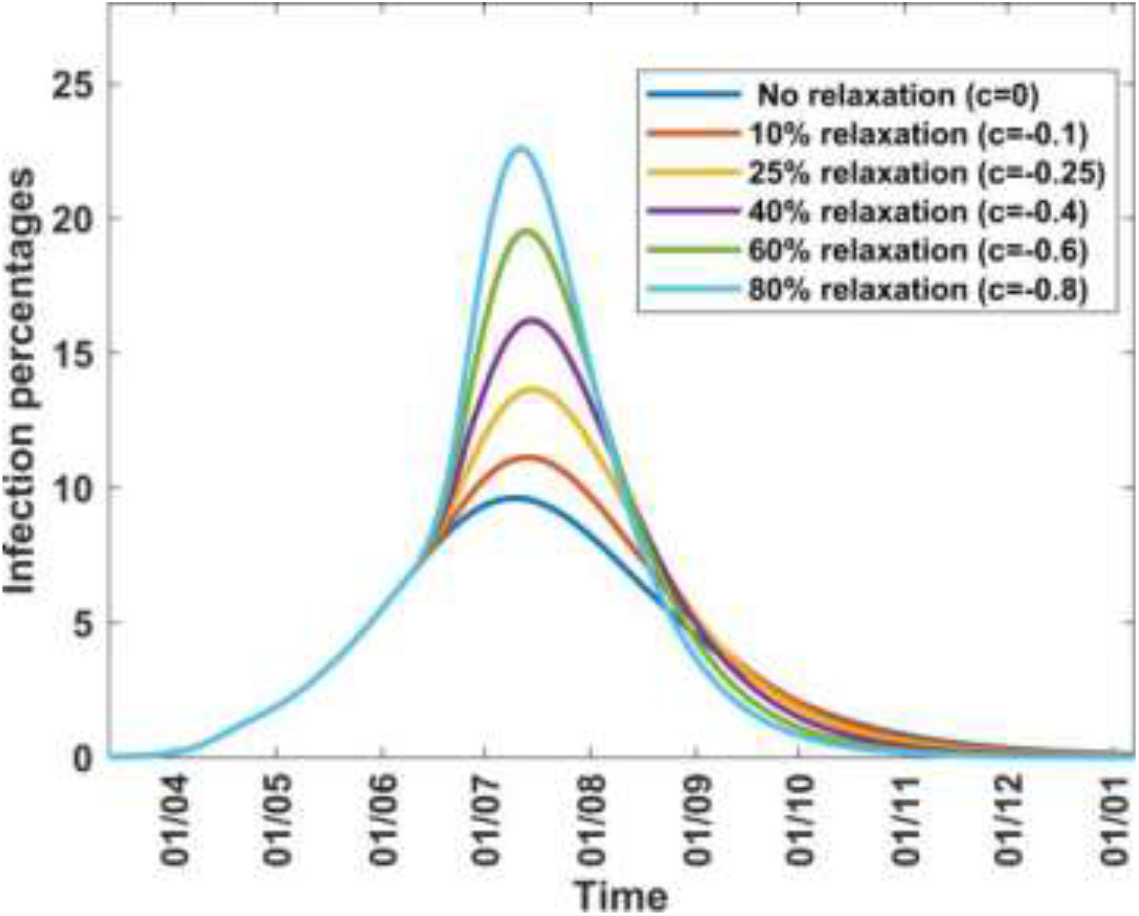
Infection percentages for various relaxation scenarios on 40% mitigation.

Since the relaxation is on the 40% mitigation of the baseline dynamics, the transmission rate up to the time of the relaxation, *β*_*b*_(*t*), must be the mitigation transmission corresponding to c = 0.4 in Table 4, namely, *β*_*b*_(*t*) = 0.1107. Using this value of *β*_*b*_(*t*) in Equation (14), with m = 15, and drawing the curves associated with different values of c, we obtain Figure 6. For a given value of c the curve shows the trajectory of the infection as a result of a one-time relaxation force imposed, on 8^th^ June 2020, upon a 40% mitigation force, with no other subsequent interventions. It is seen that the infection peak increases with decrease in c, or increase in its magnitude, and relaxation percentage.

Table 5 shows how varying the parameter c affects the transmission rate, the effective reproduction number, from Equation (16), and the infection. It shows that continued decrease in c, or increase in relaxation percentage, leads to a situation where the effective reproduction number becomes 2.99 (for c = −0.8 or 80% relaxation) and thus exceeds the basic reproduction number of 2.76. This reflects the fact that rapid lifting of mitigation measures can result in an outbreak of faster spreading covid-19, as has been reported in several countries since the outbreak of the disease. Public health advice is that mitigation measures should not be relaxed too rapidly. Table 5 also gives the infection at the end of the relaxation period, namely 8th August 2020; it enables the planner, who on 8th June 2020 is proposing a 2-month relaxation action, to forecast the infection percent on 8th August 2020, depending on the force of the relaxation.

**Table 5:** Scenarios involving changes in the parameter c under relaxation. The following values are used *β*_*b*_ = 0.1107, *R*_*b*_ = 1.660, *γ* = 0.0518 and *δ* = 0.015.

| $c$ | % change<br>in $\beta_b$ | $\beta$ | $R_e$ | % peak<br>infection | % infection<br>8 August |
| --- | --- | --- | --- | --- | --- |
| 0 | 0 | 0.1107 | 1.66 | 9.6 | 6.6 |
| -0.1 | 10 | 0.1218 | 1.82 | 11.1 | 7.7 |
| -0.25 | 25 | 0.1384 | 2.07 | 13.6 | 8.8 |
| -0.4 | 40 | 0.1550 | 2.32 | 16.2 | 9.2 |
| -0.6 | 60 | 0.1771 | 2.65 | 19.5 | 9.1 |
| -0.8 | 80 | 0.1993 | 2.99 | 22.6 | 8.5 |

A question similar to what was raised in the mitigation assessment is the following: Can we quantify the impact of the relaxation measures effected from 8th June to 8th August? To answer this question a priori we would have to estimate, before 8th June 2020, the value of the parameter c as described in Section 5. We can also answer this question a posteriori, after 8th August 2020, by making use of the results in Section 6.1 (c), where we solved the system in Equation (14) from the onset of the disease, namely 13^th^ March 2020, through the mitigation period, till the end of the relaxation period, namely, 8^th^ August 2020. Table 2 shows that from the end of the mitigation period to the end of the relaxation period, the effective reproduction number increases from 1.66 to 2.07. This change represents an increase of approximately 25% between the two periods and hence it is logical to take *c* = *−*0.25 as the relaxation parameter in Equation (14) and the trajectory of the infection would be the curve identified by 25% relaxation in Figure 6.

##### (c) Combination of mitigation and relaxation effects

To assess the effect of interventions during the first wave of COVID-19 in Kenya, we trace the trajectory of infection, taking into consideration the impact of the mitigation and relaxation measures implemented by the Kenya government. The trajectory consists of three distinct parts as indicated hereafter.

###### Baseline trajectory

This consists of the curve labelled “No mitigation (c=0)” in Figure 5. This curve is reproduced in Figure 7 (black curve) and is shown in two parts: the continuous portion, from 13th March to 8th April 2020 shows the percent infection trajectory during the baseline period and the dotted portion, after 8th April, indicates the projected trajectory in the absence of further interventions. As we shall see later, this projected trajectory is not followed due to subsequent mitigation and relaxation actions.

**Figure 7:**
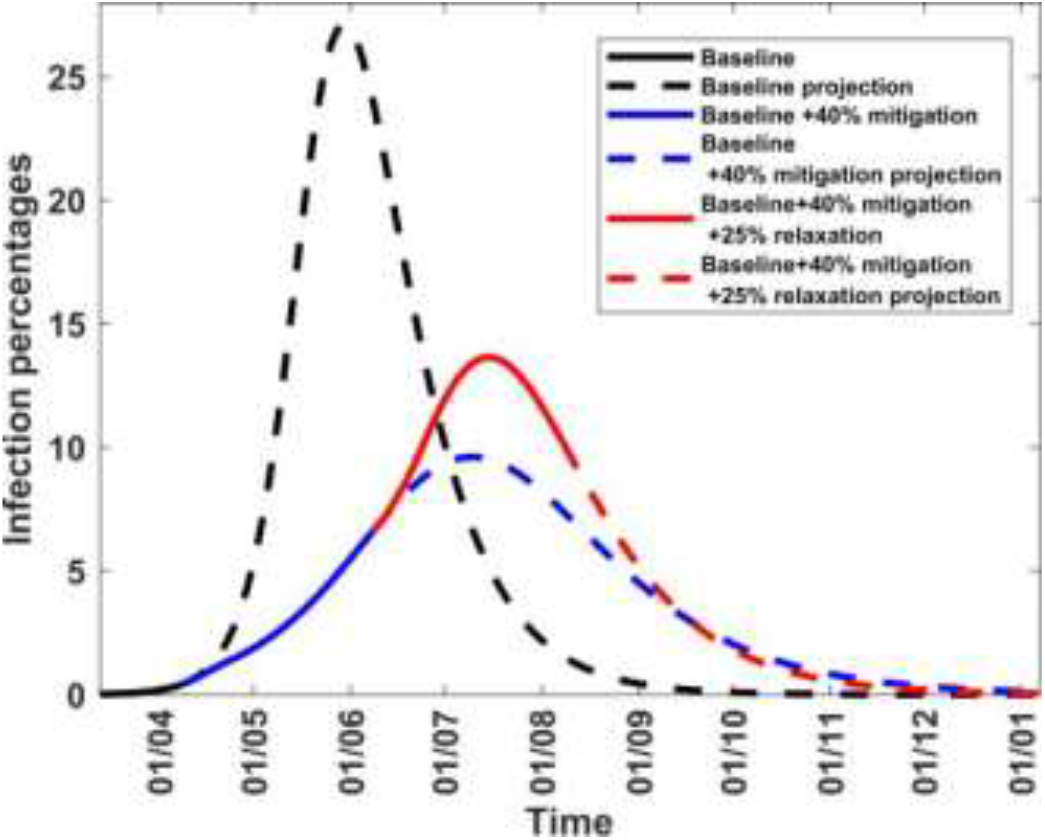
Infection percentages for different mitigation or relaxation dynamics. The red curve shows infection trajectory arising from 40% mitigation followed by 25% relaxation, the blue curve shows the infection trajectory arising from mitigation dynamics while the black curve shows infection trajectory arising from baseline dynamics.

###### Mitigation trajectory

This consists of the curve labelled “40% mitigation (c=0.4)” in Figure 5. This curve is reproduced in Figure 7 (blue curve) and is shown in two parts: the continuous portion, from 9th April to 8th June 2020 shows the percent infection trajectory during the mitigation period and the dotted portion, after 8th June, indicates the projected trajectory in the absence of further interventions. We see later that this projected trajectory is also not followed due to subsequent relaxation action.

###### Relaxation trajectory

This consists of the curve labelled “25% relaxation (c=-0.25)” in Figure 6. This curve is reproduced in Figure 7 (red curve) and is shown in two parts: the continuous portion, from 9th June to 8th August 2020 shows the percent infection trajectory during the relaxation period and the dotted portion, after 8th August, indicates the projected trajectory in the absence of further interventions. We see later that this projected trajectory is followed until the second COVID-19 wave emerges in early September 2020.

###### Combined trajectory

Combining the continuous portions of the curves in Figure 7, yields the trajectory of the infection from the onset of the disease to the end of the relaxation period, namely 8th August 2020.. The result is in Figure 8, where the 7-day moving averages are given and are compared with observed values. The projected trajectory is extended to the beginning of the 2nd wave in early September, for explanations given in the preceding paragraph. It can be seen that the predicted percent infection trajectory reasonably matches the observed values during the first wave of COVID-19 till early September when the second wave commences. The extension of the projected trajectory beyond the beginning of the 2nd wave indicates that had there not been further events that contributed to a second wave surge, COVID-19 infections would have reduced to insignificant levels by January 2021.

**Figure 8:**
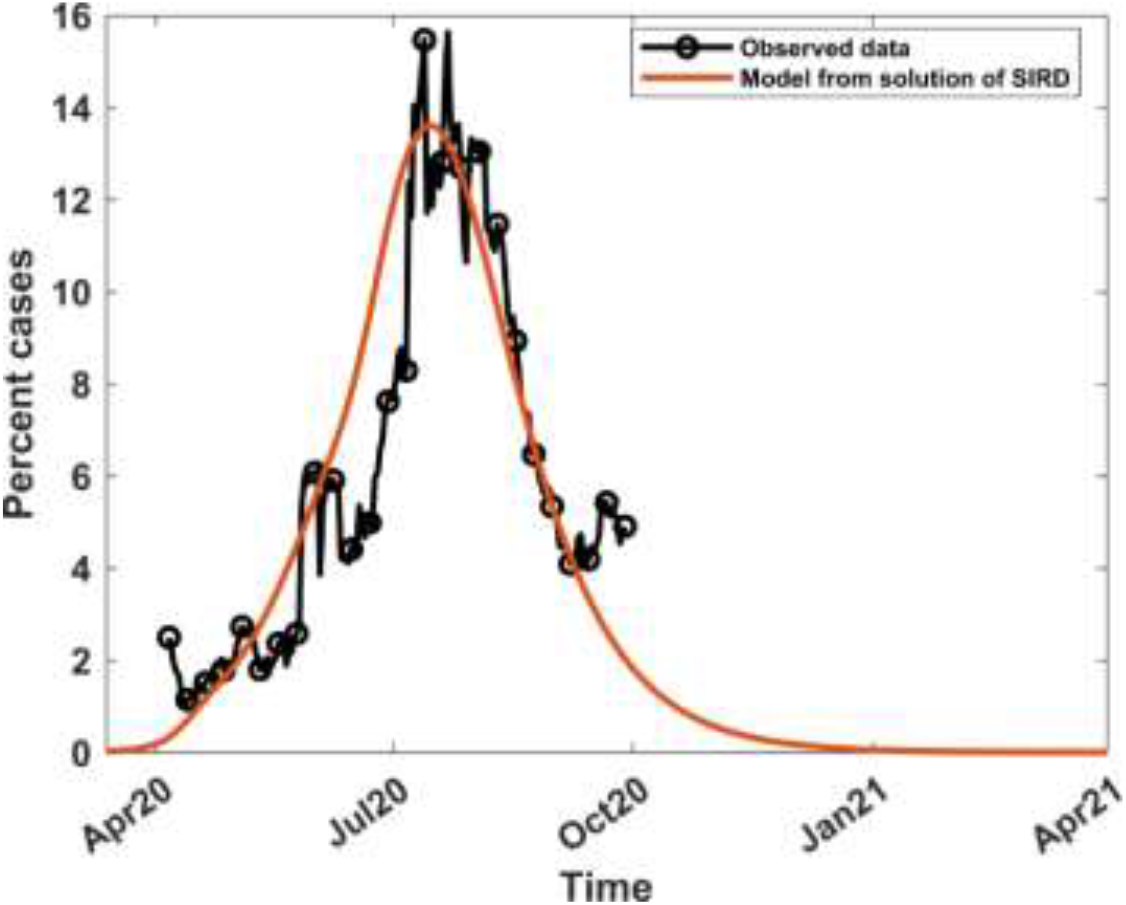
The 7-day moving average of observed and computed percent infections combining mitigation and relaxation strategies.

#### 6.2.2 Determination of intervention magnitudes given positivity rates

In this section, the observed positivity curve is given and we seek to estimate the mitigation and relaxation values that could have yielded such a curve. The problem can be solved only *a posteriori*, and it will enable assessment of the interaction between interventions and disease dynamics. The approach involves generating a suitable surface envelope around the curve and using the trajectory of the midpoint of the envelope to reflect the computed positivity curve. The concept of envelopes is applied to numerous phenomena, including robotics and kinematics, motion involving collision avoidance, gear transmission design [58 – 60]. To start the process, we require a suitable initial surface around the curve that is not too large, for computational efficiency. The surface is bounded by curves defined by constant values of mitigation and relaxation percentages. We have already seen that hard lockdown was defined as mitigation of 44% and moderate lockdown as mitigation of 25% [23]; it is therefore reasonable to start with mitigation percentages in the range (25, 50). Relaxation techniques applied after mitigation are designed to increase the transmission rate, preferably to 80% - 90% of the pre-mitigation transmission. If we assume that the relaxation increases the transmission to 90% of the pre-mitigation level then from Equation (14), we conclude that relaxation percentages will be in the range (20, 80). To identify the initial width of the envelope, we choose a fixed relaxation percentage and draw curves of interventions consisting of mitigation values in the range (25, 50), or a suitable subset of it, followed by the selected relaxation. The curves all start at the same point but, as time progresses, they begin to diverge till two curves appear on either side of the observed positivity rate; this enables us to define the initial left and right boundaries of the envelope. Further configurations involving intermediate mitigation percentages and other relaxation percentages can be made, if necessary, to enable identification of an initial envelope that is closer to the positivity rate curve.

To generate the envelope, we considered the time series values of infection percentages obtained from mitigation and relaxation percentages selected above. From these time series, maximum and minimum infection percentages are obtained at each time point and they are taken as the boundaries of the envelope, as shown in the final optimized envelope in Figure 9. The midpoint of the envelope trajectory, shown in red, reflects the computed positivity rate curve, with a mitigation of 42.5% and relaxation of 26.0% as averaged from values at the midpoints of the family of envelopes. Solution of the SIRD model in Section 6.1 yielded mitigation of 40% and relaxation of 25%. Although the approach differed significantly from the current one, the two methods yield intervention parameters with less than 10% relative error from each other and illustrates significant consistency in the findings.

**Figure 9.**
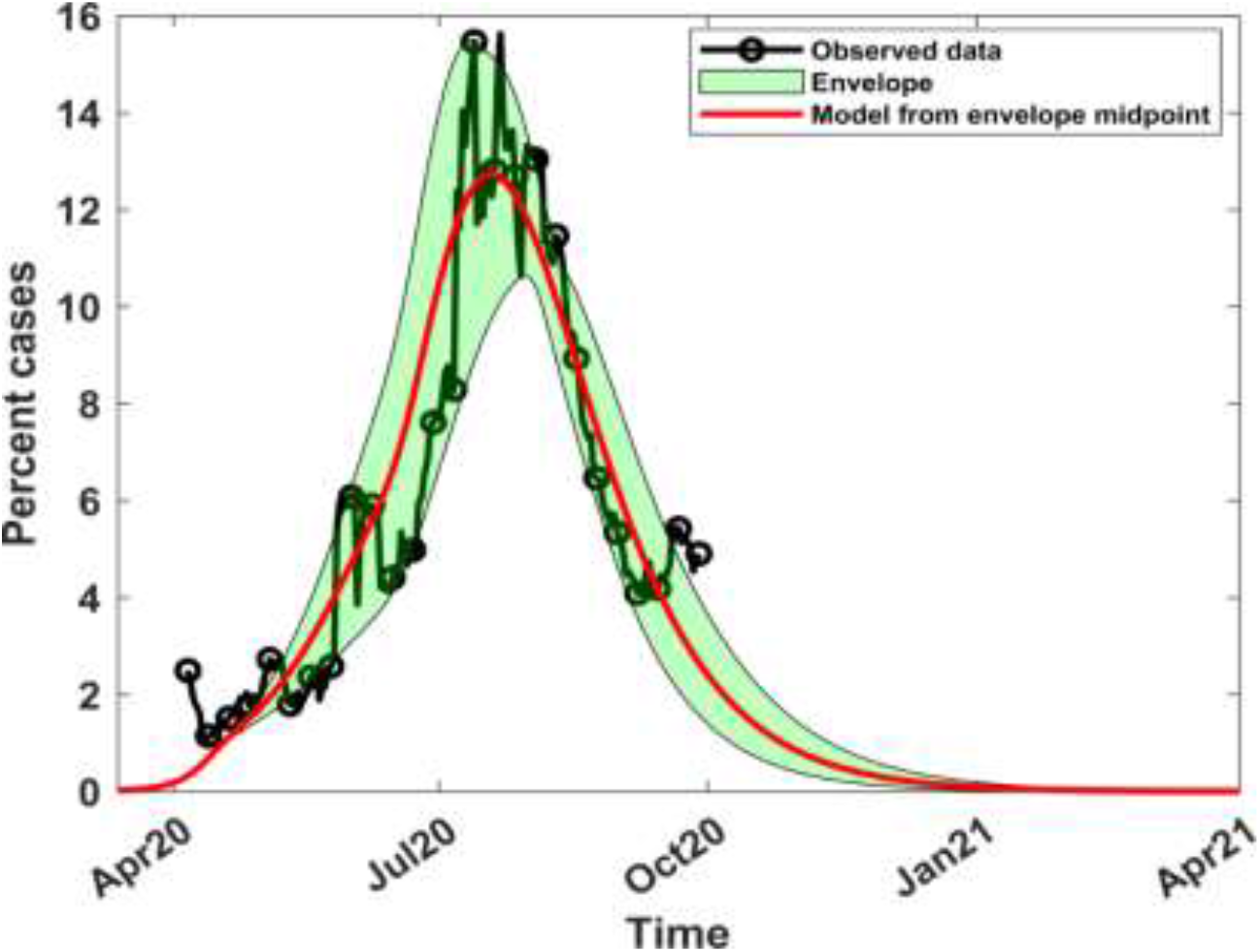
Optimized envelope around the positivity rate curve, with computed positivity curves from the envelope midpoint

## 7 Conclusions

We have developed a new numerical scheme for determining initial estimates of the COVID-19 parameters in the SIRD model. The parameters can be refined by a simple search of the minimum point, instead of more complex procedures. The results yield values of R_0_ which are comparable with other numerical schemes, thus validating our approach. We focused on the case where the death rate was known and there was a need to estimate only the recovery rate; the method can, however, be extended readily to estimate the death and recovery rates. By carrying out computations from the onset of the disease to the end of intervals that coincide with major intervention measures, we can quantify the effect of the interventions. There is a need for further analysis to determine the optimum grid outlay.

The mathematical model for interventions takes into account mitigation, which results in transmission rate decrease and in relaxation, which results in a transmission rate increase, hence in spikes. The model yields an infection curve, subject to intervention measures, that closely follows the observed trends. The process depends on a parameter, whose value is positive for mitigation and negative for relaxation. This parameter should be computed a priori, if the method is to be used as a basis for decision-making in order to enact guidelines commensurate with the level of planned interventions. The computation would require a multidisciplinary team of researchers drawn from diverse backgrounds. In determining a surge in the transmission, it should not be assumed that the spike is purely a result of the government having lifted some mitigation measures. It is often a result of society violating the laid down mitigation guidelines and, in some cases, openly protesting and against lockdowns as a result of “COVID-fatigue”. Further investigation should be carried out to determine how the method here, and that in the previous paragraph, can be extended to second and subsequent waves.

## Data Availability

Data sources: All the data used is in the public domain [1, 44, 56, 57]

## Acknowledgements

We acknowledge Alice Wangui Wachira, Anne Kinyua and Lucy Nyanchama for their assistance with data collection.. Victor Juma acknowledges the support from KEMRI-Wellcome Trust Research Programme (KEMRI-WTRP)

## Data sources

All the data used is in the public domain [1, 44, 56, 57]

## Notes

### Competing Interest Statement

The authors have declared no competing interest.

### Clinical Trial

Not a clinical trial

### Funding Statement

No funding support

### Author Declarations

KNH-UoN Ethics and Research Committee https://erc.uonbi.ac.ke

## References

[1]. Worldometer. https://www.worldometers.info/coronavirus/ Accessed on diverse dates from March 2020 to February 2021.

[2]. Africa CDC: Centres for Disease Control and Prevention. www.africacdc.org/covid-19/, August 2020.

[3]. Coronaviridae Study Group of the International Committee on Taxonomy of Viruses. The species severe acute respiratory syndrome-related coronavirus: classifying 2019-nCOV and naming it SARS-CoV-2. Nat. Microbiology, 2020, 5(4): 536–544.

[4]. Lanying Du, Yuxian He, Yusen Zhou, Shuwen Liu, Bo-Jian Zheng, and Shibo Jiang. The spike protein of SARS-CoV-a target for vaccine and therapeutic development. Nature Reviews Microbiology, 2009, 7(3):226–236.

[5]. Nicole M. Bouvier and Peter Palese.The biology of influenza viruses, Vaccine, 2008 Sep 12; 26(Suppl 4); D49–D53.

[6]. Yushun Wan, Jian Shang, Rachel Graham, Ralph S Baric, and Fang Li. Receptor recognition by the novel coronavirus from Wuhan: An analysis based on decade-long structural studies of SARS Coronavirus. Journal of virology, 2020, 94(7), [e0012720]. https://doi.org/10.1128/JVI.--27-20.

[7]. World Health Organization. Modes of transmission of virus causing COVID-19: implications for IPC precaution recommendations: scientific brief, 27 March 2020. World Health Organization. https://apps.who.int/iris/handle/10665/331601. License: CC BY-NC-SA 3.0 IGO, 2020.

[8]. World Health Organization. Report of the WHO-China Joint Mission on Coronavirus Disease 2019 (COVID-19), 16 – 24 February 2020.

[9]. Yousef Alimohamadi, Maryam Taghdir, and Mojtaba Sepandi. The estimate of the basic repro-duction number for novel Coronavirus disease (COVID-19): a systematic review and meta-analysis. J Prev Med Public Health. 2020 May; 53(3):151–157.DOI:10.3961/jpmph.20.076.

[10]. Ying Liu, Albert A Gayle, Annelies Wilder-Smith, and Joacim Rocklöv. The reproductive number of COVID-19 is higher compared to SARS coronavirus. J Travel Med. 2020 Mar 13; 27(2):taaa021. DOI:10.1093/jtm/taaa021, 2020.

[11]. Mathew Biggerstaff, Simon Cauchemez, Carrie Reed et al. Estimates of the reproduction number for seasonal, pandemic, and zoonotic influenza: a systematic review of literature. BMC Infec. Dis 14, 480 (2014). https://doi.org/10.1186/1471-2334-14-480., 2014.

[12]. ASM. https://asm.org/Articles/2020/July/COVID-19-and-the-Flu [accessed 11 February 2021]

[13]. CDC. https://www.cdc.gov/flu/symptoms/flu-vs-covid19.htm [accessed 11 February 2021]

[14]. Simon Thornley and Arthur J. Morris. Rapid Response to “The COVID-19 Elimination debate needs correct data”, BMJ 2020; 371:m3883, 2020.

[15]. Fernando Javier Aguilar Canto and Eric José Avila-Vales. Fitting parameters of SEIR and SIRD models of COVID-19 pandemic in Mexico. ResearchGate. 2020.

[16]. Aniruddha Adiga, Devdatt Dubhashi, Bryan Lewis, Madhav Marathe, Srinivasan Venkatramanan & Ani Vullikanti. Mathematical models for COVID-19 pandemic: A comparative analysis. Journal of the Indian Institute of Science 100, 793–807 (2020). https://doi.org/10.1007/s41745-020-00200-6

[17]. Joel Hellewell, Sam Abbott, Amy Gimma, Nikos I Bosse, Christopher I Jarvis, Timothy W Russell, James D Munday, Adam J Kucharski, W John Edmunds, Fiona Sun, et al. Feasibility of controlling COVID-19 outbreaks by isolation of cases and contacts. The Lancet Global Health, Volume 8, Issue 4, E488–E496, April 01, 2020.

[18]. Kentaro Iwata and Chisato Miyakoshi. A simulation on potential secondary spread of novel coronavirus in an exported country using a stochastic epidemic SEIR model. J Clin. Med. 2020 9(4):944; https://doi.org/10.3390/jcm9040944, 2020.

[19]. Ruiyun Li, Sen Pei, Bin Chen, Yimeng Song, Tao Zhang, Wan Yang, and Jeffrey Shaman. Substantial undocumented infection facilitates the rapid dissemination of novel coronavirus (SARS-CoV-2). Science, 368(6490):489–493, 2020. DOI:10.1126/science.abb3221.

[20]. PHP Cintra, MF Citeli, and FN Fontinele. Mathematical models for describing and predicting the COVID-19 pandemic crisis. arXiv:2006.02507v, 2020.

[21]. Elena Loli Piccolomini and Fabiana Zama (2020) Monitoring Italian COVID-19 spread by a forced SEIRD model. PLoS ONE, 15(8):e0237417. https://doi.org/10.1371/journal.pone.02347417

[22]. Ivan Korolev. Identification and estimation of the SEIRD epidemic model for COVID-19. J. Econom 220(1): 63–85, 2021. https://doi.org/10.1016/j.econom.2020.07.038.

[23]. Isabel Frost, Gilbert Osena, et al. Modelling COVID-19 transmission in Africa: countrywise projections of total and severe infections under different lockdown scenarios. BMJ Open 2021;11:e044149. doi:10.1136/bmjopen-2020-044149.

[24]. Jesús Fernández-Villaverde and Charles I Jones. Estimating and simulating a SIRD model of COVID-19 for many countries, states, and cities. National Bureau of Economic Research, Working Paper 27128, DOI 10.3386/w27128. May 2020.

[25]. Cleo Anastassopoulou, Lucia Russo, Athanasios Tsakris, and Constantinos Siettos (2020) Data-based analysis, modelling and forecasting of the COVID-19 outbreak. PloS ONE, 15(3):e0230405. https://doi.org/10.1371/jounal.pone.0230405.

[26]. Diego Caccavo. Chinese and Italian covid-19 outbreaks can be correctly described by a modified SIRD model. medRxiv 2020, https://doi.org/10.1101/2020.03.19.20039388.

[27]. Ian Cooper, Argha Mndal and Chris G. Antonopoulos. A SIR model assumption for the spread of COVID-19 in different communities. Chaos, Solitons and Fractals, 2020 Oct; 139:110057. doi: 10.1016/j.chaos.2020.110057.

[28]. Enahoro A Iboi, Oluwaseun O Sharomi, Calistus N Ngonghala, and Abba B Gumel. Mathematical Modeling and Analysis of COVID-19 pandemic in Nigeria. Mathematical Biosciences and Engineering,2020, 17(6): 7192–7220. doi: 10.3934/mbe.2020369..

[29]. Samuel Mwalili, Mark Kimanthi, Viona Ojiambo, Duncan Gathungu, and Rachel Waema Mbogo. SEIR model for COVID-19 dynamics incorporating the environment and social distancing. BMC Research Notes 13, 352 (2020). https://doi:10.1186/s13104-020-05192-1.

[30]. Jan-Diederik van Wees, Sander Osinga, Martijn van der Kuip, Michael Tanck, Maurice Hanegraf, Marten Pluymaekers, et. al. Forecasting hospitalization and ICU rates of the COVID-19 outbreak: an efficient SEIR model. [Preprint]. Bull. World Health Organ., E-pub:30 March 2020.

[31]. Masaki Tomachi and Mitsuo Kono. A mathematical model for COVID-19 pandemic – SIIR model: Effects of asymptomatic Individuals. Journal of General and Family Medicine, Volume 22, Issue 1, Pages 5–14, January 2021.

[32]. Mustapha Serhani and Hanane Labbardi. Mathematical modelling of COVID-19 spreading with asymptotic infected and interacting peoples. J. Appl. Math. Comput. (2020). https://doi.org/10.1007/s12190-020-01421-9, 2020.

[33]. Barradas Ignacio, Hugo Flores Arguedas, Ariel Camacho and Fernando Saldanã. Modeling the transmission dynamics and the impact of the control interventions for the COVID-19 epidemic outbreak. Mathematical Biosciences and Engineering, 2020, 17(4): 4165–4183. doi: 10.334/mbe.2020231.

[34]. Guilherme S. Costa, Wesley Cota and Silvio C. Ferreira. Metapopulation modelling of COVID-19 advancing into countryside: an analysis of mitigation strategies for Brazil. medRxiv 2020. https://doi.org/10.1101/2020.05.06.20093492.

[35]. Davide Faranda and Tommaso Alberti. Modeling the second wave of COVID-19 infections in France and Italy via stochastic SEIR model. Chaos, Vol,30, 111101 (2020); https://doi.org/10.1063/5.0015943.

[36]. Sansao A. Pedro, Frank T. Ndjomatchoua, Peter Jentsch, Jean Tchuenche, Madhur Anand and Chris T. Bauch. Conditions for a second wave of COVID-19 due to interactions between disease dynamics and social processes. Front. Phys. 09 October 2020, https://doi.org/10.3389/fphy 2020.574514.

[37]. Efthimios Kaxiras and Georgios Neofotistos. Multiple epidemic wave model of the COVID-19 pandemic: Modeling Study, J. Med Internet Res. 2020 Jul: 22(7): e20912.

[38]. Giacomo Cacciapaglia and Francesco Sannino. Second wave COVID-19 pandemics in Europe: a temporal playbook. Scientific Reports, 10, Article number: 15514 (2020).

[39]. Steffen E Eikenberry, Marina Mancuso, Enahoro Iboi, Tin Phan, Keenan Eikenberry, Yang Kuang, Eric Kostelich, and Abba B Gumel. To mask or not to mask: Modeling the potential for face mask use by the general public to curtail the COVID-19 pandemic. Infect. Dis Model. 2020;5:293–308.

[40]. Kiesha Prem, Yang Liu, Timothy W. Russell, Adam J. Kucharski, Rosalind M. Eggo, Nicholas Davies, et.al. The effect of control strategies to reduce social mixing on outcomes of the COVID-19 epidemic in Wuhan, China: a modelling study. The Lancet. Vol. 5, Issue 5, E 261–270, May 01, 2020. DOI: https://doi.org/10.1016/S2468-2667(20)30073-6.

[41]. Sara MA S Elsheikh, Mohamed K Abbas, Mohamed A Bakheet, and AM Degoot. A mathematical model for the transmission of coronavirus disease (COVID-19) in Sudan. ResearchGate. DOI:10.13140/RG.2.2.24167.27043/1, May 2020.

[42]. Solym Manou-abi and Julien Balicchi. Analysis of the COVID-19 epidemic in French overseas department Mayotte based on a modified deterministic and stochastic SEIR model. MedRxiv 2020. https://doi.org/10.101/2020.04.15.20062752.

[43]. First case of coronavirus disease confirmed in Kenya. https://www.health.go.ke/first-case-of-coronavirus-disease-confirmed-in-kenya/. [accessed 18th December 2020]

[44]. Ministry of Health, Kenya. https://www.health.go.ke/ [accessed on diverse dates from March 2020 to February 2021]

[45]. Kenya Gazette Supplement No. 41. The Public Health Act (Cap.242). the public health (COVID-19 restriction of movement of persons and related measures) rules. 16^th^ April 2020.

[46]. Presidential addresses on COVID-19. https://www.president.go.ke [accessed on diverse dates from March 2020 to February 2021]

[47]. Academia Kenya. www.academia-ke.org/library/tag/covid19/. [accessed on diverse dates from March 2020 to February 2021].

[48]. Gerardo Chowell, Cécile Viboud, Lone Simonsen, Mark Miller, and Wladimir J Alonso. The reproduction number of seasonal influenza epidemics in Brazil, 1996–2006. Proc Biol Sci. 2010 Jun 22: 277(1689):1857–1866..

[49]. Ka Chong, William Goggins, Benny Zee, and Maggie Wang. Identifying meteorological drivers for the seasonal variations of influenza infections in a subtropical city—Hong Kong. Int J Environ Res Public Health. 2015 an 28;12(2):1560–1576. DOI:10.3390/ijerph120201560.

[50]. Victor Ogesa Juma. Modelling influenza incidence in relation to meteorological parameters in Kenya. 2015.

[51]. Cesar Lopez. MATLAB Optimization Techniques. Apress, 2014.

[52]. William E Hart, Carl D Laird, Jean-Paul Watson, David L Woodruff, Gabriel A Hackebeil, Bethany L Nicholson, and John D Siirola. Pyomo-Optimization Modeling in Python. Springer, 2017.

[53]. Paulo Cortez. Modern Optimization with R. Springer-Verlag, 2014.

[54]. Daisuke Furushima, Shoko Kawano, Yuko Ohno, and Masayuki Kakehashi. Estimation of the Basic Reproduction Number of Novel Influenza A (H1N1) pdm09 in Elementary Schools Using the SIR Model. Open Nurs J. 2017 Jun 29;11:64–72. doi: 10.2174/1874434601711010064

[55]. Tianfeng Chai and Roland R Draxler. Root mean square error (RMSE) or mea.n absolute error (MAE)?–Arguments against avoiding RMSE in the literature. Geosci. Model Dev.,7, 1247–1250, 2014, https://doi.org/10.5194/gmd-7-1247-1250.

[56]. World Health Organisation. https://www.who.int/emergencies/diseases/novel-coronavirus-2019. [accessed on diverse dates from March 2020 to February 2021]

[57]. Our World in Data. https://www.ourworldindata.org/coronavirus. [accessed on diverse dates from March 2020 to February 2021]

[58]. Helmut Pottmann and Martin Peternell. Envelopes – Computational theory and applications. In: Spring Conference on Computer Graphics, pp.3-23. Comenius University, Bratislava, 2000.

[59]. Martina Batorova and Pavel Chalmoviansky. Envelopes of planar curves in applications. http://www.lepageri.eu/files/Chalmoviansky-prezntacia.pdf. [accessed 2nd March 2021].

[60]. Thierry Dana-Picard and Nurit Zehavi. Revival of a classical topic in Differential Geometry: the exploration of envelopes in a computerized environment. International Journal of Mathematical Education in Science and Technology, 47:6, 938–959, DOI:10.1080/0020739X.2015.1133852.

